# Mass-Standardised Antibody Affinity Maturation to the Spike Protein of SARS-CoV-2 Omicron Variants in a Constant-Exposure Cohort: forgiving Original Antigenic Sin

**DOI:** 10.1101/2022.12.01.22282932

**Authors:** Philip H. James-Pemberton, Shivali Kohli, Aaron C. Westlake, Alex Antill, Rouslan V. Olkhov, Andrew M. Shaw

## Abstract

Prevalence of SARS-CoV-2 amongst school children in the omicron wave was high leading to a constant exposure of teachers to the virus. A cohort of staff members (*n* = 28) was recruited from a UK primary school setting. The prevalent variants at the time were Omicron BA.1, BA.4/5 and BA.2: 61% of the cohort reported a lateral-flow-confirmed positive test for SARS-CoV-2 infection in late 2021 or 2022. A mass-standardised quantitative antibody spectrum was measured for both antibody concentration and a new measure of antibody quality; the proportion of antibodies still bound to the epitope at pH 3.2 compared with pH 7.4. The antibody spectrum was measured or IgG to the spike protein for variants Wuhan, Alpha, Beta, Gamma, Delta and Omicron BA.1, BA.2.75, BA.2.12.1, BA.4 and BA.5. The new antibody quality measure shows a mean of 49% with (IQR 32% – 68%). The cohort showed a Universal positive immunity endotype, U(+), of antibodies binding to the spike protein across the variant spectrum with an incidence of 78% (95% CI 60% – 88%) with good antibody concentrations to all ten variants; the incidence drops to 25% (95% CI 13% – 43%) when the affinity spectrum is measured.

## Introduction

School staff members are uniquely exposed to transient infections in children when in enclosed, poorly ventilated environments, including during the SARS-CoV-2 pandemic^1^. A retrospective test-and-trace study from Italy^2^ has shown that, of the school students in the age range 6 –10 years, 50% ( 95% CI, 39% – 60 %) tested positive and were asymptomatic carriers and 21.4% (95% CI 14% – 31%) had symptomatic infections for SARS CoV-2. Similarly, studies in the UK following the release of school teaching restrictions showed a surge in student infections during the period August to October 2021^3^ with household clusters adding to transmissions in parents, particularly from children aged 5-15. Other studies counter with findings that school children in the pandemic^4^ in the age range 5–11 years old did not have a prevalence of infection greater than the local working population, although the prevalence of asymptomatic carriers was not reported.

A teachers’ cohort is a constant-exposure cohort addressing directly the concepts of immunity imprinting and original antigenic sin ^5–7^ with continual challenges evolving the antibody epitope selection and the T cell response. In a previous study^8^, we observed antibody immunity endotypes to a spectrum of variants: Wuhan, Alpha, Beta, Gamma, Delta and Omicron variants: BA.1, BA.4, BA.5, BA.2.75 and BA.2.12.1. A universal endotype showed antibody concentrations to all variants was present in 75% of the cohorts tested. The universal protection is consistent with raising antibodies to conserved epitopes, such as the hinge region required for the Spike protein to transition from pre-fusion to fusion state^9^. Meanwhile, immunity endotypes of the remaining 25% show poor antibody production or drop-outs to one or more of the variants, having made antibodies to epitopes that are not present in all variants, potentially leading to a risk of post-viral sequalae^10–12^.

Immunity endotypes were previously explored for the concentration of antibodies against variant spike proteins^8^ but affinity of the antibody-antigen interactions is also an important and potentially evolving immunity endotype and critical to the clone selection process^13^. Measurement of antibody affinity (or avidity) requires the association and dissociation rates of the paratope-epitopes complex to be accurately measured, and for stable complexes this corresponds to a half-life of several hours^14^. A high-affinity antibody in an immune response suggests the clearance proves needs a stable complex. Further, affinity studies are usually performed at pH 7 which is poor mimic of the mucosal environment where the pH may be more acidic, pH 5.5. Here, we have explored a new method of assessing the stability of the paratope-epitope complex by measuring a ratio between antibody-antigen binding responses at two different pH values: pH 3.2 and pH 7.4. Titration of the paratope-epitope complex shows that affinity (or avidity) could be pH dependent and so environment dependent. The critical parameter in forming a high-quality antibody response is the antibody-antigen complex lifetime in all environments which controls the timescales on which an antibody-mediated response such as endocytosis or complement action takes place leading to a sterilising event and removal of the virus.

In this paper, we report a prospective cohort study on 28 staff members in a primary school setting with samples taken 13^th^ July 2022. The local prevalent Omicron variants at the time of sampling were BA.4 and BA.5, whilst BA.1.1 and BA.2 were prevalent in the preceding six months. Demographic data was collected alongside vaccination histories and lateral flow confirmations of any recent infections. A COVID antibody immunity test was performed for nucleocapsid, Spike (Wuhan), Spike Omicron BA.1 and receptor binding domain. Further, concentration and pH-dependent affinity profiles were collected for the spike proteins of the variants Wuhan, Alpha, Beta, Gamma, Delta, as well as Omicron strains BA.1, BA.2.75, BA.2.12.1, BA.4 and BA.5. The full 20-dimensional profile of concentration and pH-dependent affinity for 10 variants for each adult tested in the school was interpreted in the constant-exposure cohort context and optimisation of the high-avidity antibody response.

## Materials and Methods

### Materials

Materials used throughout the course of the experiments were used as supplied by the manufacturer, without further purification. Sigma-Aldrich supplied phosphate buffered saline (PBS) in tablet form (Sigma, P4417), phosphoric acid solution (85 ± 1 wt. % in water, Sigma, 345245) and Tween 20 (Sigma, P1379). Glycine (analytical grade, G/0800/48) was provided by Fisher Scientific. Assay running and dilution buffer was PBS with 0.005 v/v % Tween 20 and the regeneration buffer was 0.1 M phosphoric acid with 0.02 M glycine solution in deionized water.

A recombinant human antibody to the spike protein S2 subdomain, a chimeric monoclonal antibody (SinoBiological, 40590-D001, Lot HA14AP2901) was used as a calibrant. The antibody was raised against recombinant SARS-CoV-2 / 2019-nCoV Spike S2 ECD protein (SinoBiological, 40590-V08B). This was selected as it could bind to all nine Spike protein variants used herein.

NISTmAb, a recombinant humanized IgG1ĸ with a known sequence^15^ specific to the respiratory syncytial virus protein F (RSVF)^16^, was from National Institute of Standards and Technology (RM8671). The detection mixture consisted of a 200-fold dilution of IG8044 R2 from Randox in assay running buffer. The Spike Variant sensor chips were printed as detailed elsewhere^17^. The Omicron sub-variant sensor chips were printed with recombinant human serum albumin from Sigma-Aldrich (A9731), protein A/G from ThermoFisher (21186), and five SARS-CoV-2 Spike Protein variants for the Wuhan, Omicron BA.1, BA.4, BA.5, BA.2.75 and BA.2.12.1 strains from SinoBiological and BA.2.75 strain from Acro Biosystems as detailed in Table S1.

### Methods

#### Biophotonic Multiplexed Immuno-kinetic assay

The multiplexed biophotonic array platform technology has been described in detail elsewhere^8,17–20^ and has a CE mark to perform the Attomarker COVID Antibody Immunity and a UKCA mark for the Covid Antibody Spectrum Test. Briefly, the technology is a localised particle plasmon gold sensor which scatters light proportional to the mass of material (formally refractive index) near to the gold nanoparticle surface in the plasmon field. The gold nanoparticles are printed into an array of 170 spots which are then individually functionalised with the protein required. The SARS-CoV-2 antibody targets within the CE marked COVID Antibody immunity Test are nucleocapsid, receptor binding domain, Wuhan spike protein and Omicron spike protein. The inflammation marker C-reactive protein is also measured. The assay consists of a capture step, a detection step and a regeneration step with intervening wash steps. High and low controls are used to calibrate the sensor response and are repeated at regular intervals to ensure quality control^15^ during the day.

Two further arrays were fabricated. One containing Spike proteins from SARS-CoV-2 variants, as detailed elsewhere^8^, alongside a new array containing the Omicron sub-variants as listed in Table S1. The integrity of the protein samples on the surface was tested using an anti-S2 antibody (40590-D001), chosen for calibration as the corresponding epitope is present on all variants. The concentration of the antibody was calibrated against the NIST antibody RM8671, NISTmAb, a recombinant humanized IgG1ĸ with a known sequence^15^ to assure monomeric purity of the antibody. High-control and low-control samples were made using the 40590-D001 antibody and these controls were then used to quantify results from human samples in units of mg/L. Thus, the assay response is presented as a concentration of an antibody equivalent in its binding properties to the 40590-D001 control antibody. An assay common to each sensor chip was performed using the S (Wuhan) channel, with each array carrying an identical protein. There was high correlation between S channel performance on the two array types (m, c >0.92) and so the results were treated as aligned, Table S2 and Figure S2.

An endotype classification has been derived for the spike protein variant^8^ and The average of all thresholds is 1.8 mg/L (the Limit of Detection (LoD) of the technique is 0.2 mg/L) and a universal positive endotype, U(+), has the antibody concentrations for all variants above the 1.8 mg/L threshold. A single drop-out has an antibody concentration for one variant below the threshold, *e.g.* α(−), double drop-out, two variants α(−)β(−) *etc*., for all variants and these are all labelled U(±); one or more variant antibody concentrations <1.8 mg/L. A full correlation analysis for all combinations of responses was also performed from which the correlation coefficients and lines of best fit were derived. A new measure of antibody affinity has been derived comparing the antibody binding on the surface of the target antigen after a 30 s wash step at either physiological serum pH 7.4 or pH 3.2. The lower pH of 3.2 results in some dissociation of the paratope-epitope complex leading to an effective reduction of the measured antibody concentration; a measurement of the antibodies that remain strongly bound at pH 3.2 – a high-affinity concentration. A full dissociation occurs at pH 1.9; the pH used at the longer regeneration step. The antibody profile is parameterised by the total antibody concentration (AC), the high-Quality antibody concentration (HQ) and the percentage of high-quality antibodies (HQ% = HQ/AC ×100). The cohort was used to determine reference ranges for these two new immunological measures. The HQ% from the cohort was used to estimate the HQ threshold of a sterilising serum, scaling the AC threshold derived elsewhere^17^.

### Patient Cohort

The staff members were recruited from a primary school setting and took the CE marked Attomarker Antibody Immunity Test after giving informed consent with a registered healthcare professional for the use of their residual blood samples to be used in aiding understanding of the pandemic. The venous blood samples were analysed on site within 7 minutes, and the results subsequently provided to the patients. The residual vacutainer blood was transported back to the laboratory and centrifuged at 1300 rpm for 10 minutes to separate the blood constituents. The serum was collected and subsequently used for variant profile screening.

#### Ethical Approval

The use of the Attomarker clinical samples was approved by the Bioscience Research Ethics Committee, University of Exeter.

## Results

The teachers cohort recruited from the school was 86% female and 14% male: 68% had an initial AstraZeneca (AZ) two-vaccination dose, 25% Pfizer, 4% Moderna and 4% undeclared, Table 1 (and Table S4). Seven teachers in the cohort, or 25 % (95% CI 13% – 43%), were un-boosted. Each teacher had a full concentration and pH-dependent affinity profile assessment for each of the ten spike proteins. A pH-dependent affinity titration for a teacher is shown in Figure S3 for nucleocapsid, Spike protein, Omicron Spike Protein and receptor binding domain. Each of the paratope-epitope titrations show a fall in binding of approximately 50% at pH 3.2, which was selected as the differentiating pH for the HQ and HQ%, to give a good dynamic range. The defining reference range for these new parameters is derived by assuming HQ% is a clone selection property and constant for all variants. The mean and interquartile range (IQR) for HQ% was derived from Wuhan, BA.1, BA2.12.1 BA.2.75. BA.4 and BA.5 to give HQ% is 49% (IQR 32% – 68%), Table S5.

**Table 1.** Cohort demographics including vaccination profile, infection history and endotype classification for concentration and high-affinity concentration (AstraZeneca (AZ), Undeclared (U), U(+) is the universal endotype, variant dropouts α,β,ψ,ο,ο and unclassified (UC)) (Sample IDs are not known to the patient)

| Sample ID | Interval since recent positive test | Vaccine | Booster | Number of doses | Quantity Endotype (<1.8 mg/L) | High Quality Endotype (<0.9 mg/L) | High Quality Endotype (<49%) |
| --- | --- | --- | --- | --- | --- | --- | --- |
| C104 | < 4 months | AZ | U | 3 | U(+) | U(+) | 6- |
| C105 | | Pfizer | Pfizer | 3 | W$\beta\delta$ (-) | 5- | 7- |
| C106 | < 4 months | Pfizer | Pfizer | 3 | U(+) | 3- | 9- |
| C107 | < 5 months | Pfizer | | 2 | W,BA.1,$\beta\delta$ (-) | 6- | 3- |
| C108 | < 1 month | AZ | Pfizer | 3 | U(+) | 1- | 6- |
| C109 | < 5 months | AZ | Moderna | 3 | U(+) | U(+) | 0- |
| C110 | < 6 months | Pfizer |  | 2 | U(+) | 3- | 6- |
| C111 | < 1 month | AZ | Moderna | 3 | U(+) | 4- | 10- |
| C112 |  | AZ | Moderna | 3 | U(+) | 1- | 5- |
| C113 | < 1 month | U | U | 3 | U(+) | U(+) | 6- |
| C114 | < 4 months | Pfizer | Pfizer | 3 | $\beta\delta$ (-) | 5- | 4- |
| C115 | < 5 months | AZ | Pfizer | 3 | U(+) | U(+) | 4- |
| C116 | < 3 months | AZ | U | 2 | U(+) | 1- | 8- |
| C117 | < 2 months | AZ | Moderna | 3 | U(+) | U(+) | 8- |
| C118 |  | Moderna | Moderna | 3 | U(+) | 2- | 9- |
| C119 | | AZ | | 2 | $\beta\delta$ (-) | 5- | 7- |
| C120 | < 3 months | Pfizer | Pfizer | 3 | U(+) | 3- | 7- |
| C121 |  | AZ | Moderna | 3 | U(+) | U(+) | 9- |
| C122 | < 3 months | AZ |  | 2 | U(+) | 1- | 10- |
| C123 |  | AZ | Pfizer | 3 | U(+) | 4- | 9- |
| C124 | | AZ | Pfizer | 3 | $\delta$ (-) | 4- | 7- |
| C125 |  | Pfizer |  | 2 | U(+) | U(+) | 3- |
| C126 | < 1 month | AZ | Pfizer | 3 | U(+) | 1- | 2- |
| C127 | < 1 month | AZ | Moderna | 3 | U(+) | U(+) | 8- |
| C128 | < 4 months | AZ | Moderna | 3 | U(+) | 1- | 8- |
| C129 |  | AZ | Moderna | 3 | U(+) | U(+) | 9- |
| C130 | < 8 months | AZ | Pfizer | 3 | $\delta$ BA.2.75(-) | 6- | 5- |
| C131 |  | AZ |  | 2 | U(+) | U(+) | 5- |

The mean HQ% is used to covert the sterilising serum threshold total (recovering patient cohort accuracy study^17^) to derive a threshold for the sterilising serum HQ. Both thresholds are shown for some patent antibody spectra, Figure 1. The teacher cohort has a confirmed infection incidence in the preceding six months of 61% (95% CI 42% –76%) and a U(+) incidence of 78% (95% CI 60% – 88%) consistent with the incidence observed in our previous study^8^, shown in Figure 1(A). Of the remaining six teachers, five showed a Delta drop-out endotype either alone or in combination with Beta variant: the two Beta variant drop-out endotypes were associated with a Pfizer vaccination history similar to previous observations^8^. The U(+) incidence in the cohort changes for the HQ and HQ% parameters as seen for Patient 3 Figure 1 (G, H, I) where AC concentration for each variant looks strongly U(+) but HQ antibodies are much greater in the Omicron variants with an average of 49%. The total Universal Immunity endotype, U(+), incidence falls by a third to 25% (95% CI 13% – 43%). The drop-out variants (*n*=18 or 64% (95% CI 46% – 79%)) are exclusively early variant dropouts: none are Omicron variant, Table 1.

**Figure 1.**
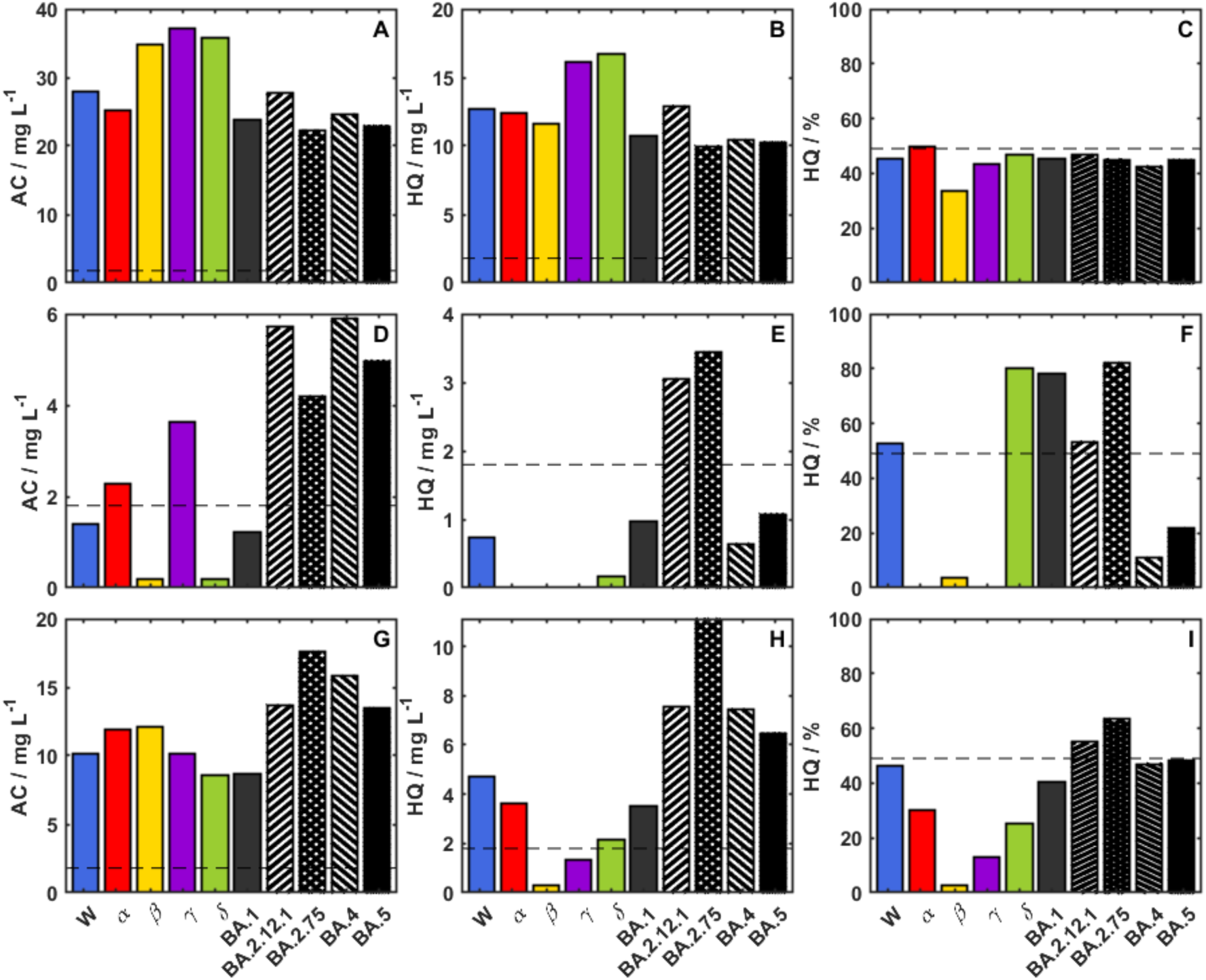
Three patients are presented, one per row, showing 10-variant antibody spectra: (A,B,C) is the profile of one patient showing antibody concentration (AC), High-quality antibodies (HQ) and the HQ as percentage of AC, HQ% – this sample is a U(+) endotype;. Patient 2 (D,E,F) shows U(−) endotype for β(−) and ο(−)for AC but falling)only BA.2.12.1 and BA.2.75 variants; and Patient 3 (G,H,I) showing a U(+) for AC but low concentrations for HQ indicating poor antibody quality except for BA2.12.1 and BA.2.75, the circulating variants. and Patient 3 (G,H,I) all show drop-out endotypes but good quality antibodies to the circulating variants at the time. The dotted line for the AC antibodies is set at the sterilising serum threshold; the dotted line for HQ is set at 49% of the sterilising threshold and the dotted line for HQ% is set at 49%.

**Figure 2.**
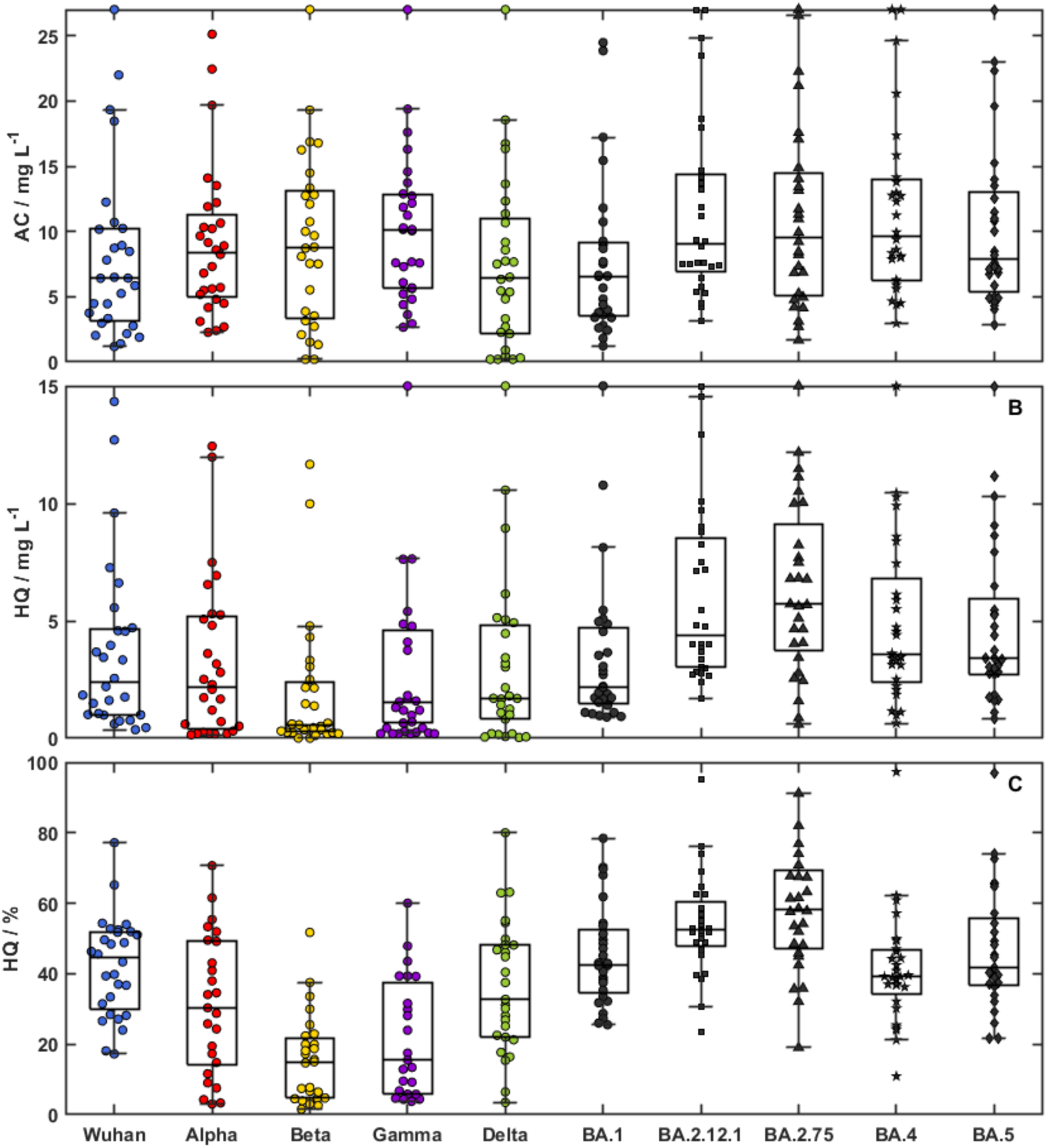
Bee-swarm plot for the Cohort Profiles: (top panel) concentration, (middle panel) high-affinity concentration and Antibody Efficiency (lower panel)

## Discussion

The teacher cohort is uniquely exposed in the school setting to children, especially of younger age, that are either asymptomatic carriers or symptomatic cases^2,3^ in the COVID pandemic. The repeated, evolving exposure is the in the ‘new-normal’ where repeated waves of infection lead to a high virus prevalence and infection. Schools have been identified^21^ as foci of the pandemic although the children themselves may not be vulnerable to infection, leading to difficult discussions about child vaccination^22^. Countries have differed approaches despite WHO recommendations^23^. The effect of re-opening schools in the UK showed significant increase in infection rates amongst school children^21^. The teachers and parents of school children represent one of the critical immunity perimeters of the pandemic with consequent implications for workforce sickness and impact on economic activity.

The constant-exposure teachers’ cohort, whether unvaccinated, vaccinated or boosted, are exposed repeatedly to the prevalent circulating virus to which there is a significantly refined immunity response. Immunity Imprinting has been reported for SARS-CoV-2 vaccination^24^ and we have reported immunity endotypes for antibody concentration against a spectrum of variant spike proteins^8^ suggesting that initial exposure to a virus or vaccine may imprint a remembered immune response. Antibody quality or antibody affinity maturation has been reported for SARS-CoV-2 immunity^25,26^ with mechanisms of maturation proposed in the germinal centre B cell response^27,28^ critical to the adaptive immunity response^29^. Sustained antigen availability has also pointed to non-standard vaccination strategies to optimise the adaptive response^30,31^. The teacher cohort is therefore an ideal setting for the observation of antibody immunity imprinting and consequence maturation: the forgiveness of original antigenic sin.

There is a spectrum of immunity in the teacher cohort derived from vaccination as well as boosted staff (four of whom reported sickness and had time off work), Table 1. The incidence of lateral-flow-test-confirmed infection in the preceding six months was 63% suggesting exposure to BA.1.159, BA.4/5 and BA.2, Figure S1. The antibody concentration profile across the spectrum of spike proteins showed the cohort incidence of U(+) was 75% and reflects previous measurements indicating that many had moderate-high antibody levels to all variants, potentially from targeting a universal epitope most likely located on the hinge region of the protein^8^. However, the new measure of antibody quality, pH-dependent affinity, shows significant variation across variants. The Wuhan antibodies generated by the boosted vaccination, and so renewed exposure to the antigen, showed affinity similar to the prevailing variants but was significantly worse for Alpha, Beta and Gamma (Kolmogorov-Smirnov test P values: Alpha=0.016, Beta=4.1×10^-8^, Gamma=1.84×10^-5^, Delta=0.28, BA.2.12.1=0.016, BA.2.75=0.0023, BA.4=0.47, BA.5=0.91, BA.1=0.91 *vs* Wuhan). Thus, the U(+) endotype incidence drops to 25%. Figure 1 (A, B, C), shows access to the universal epitope, fully affinity-matured to all variants following vaccine exposure, a highly efficient adaptive response. However, some U(+) endotypes with promising antibody concentration profiles, Figure 1 (G, H, I), do not have antibodies matured against Beta or Gamma variants, leading to lower high-affinity antibody concentration and therefore lower vaccine efficiency against these particular variants. The cohort distributions show a significant antibody maturation and consequent vaccine efficacy towards the Omicron sub-variants (High-Affinity Concentration data Kolmogorov-Smirnov test P values: Alpha=0.47, Beta=0.0023, Gamma=0.15, Delta=0.47, BA.2.12.1=0.0062, BA.2.75=0.016, BA.4/5=0.15, BA.1=0.91 *vs* Wuhan), with maximum efficiency for the variants BA.2.12.1 and BA.2.75, both descending from the widely circulated BA.2 lineage. There were no Omicron endotype dropouts in contrast to the vaccination cohort not in continuous exposure, with an incidence of 7% (95% CI 2% – 19%)^8^.

The new measure of pH-dependent affinity, HQ%, has a mean of 49% with (IQR 32% – 68%) is an easy way to assess the dissociation constant of the epitope-paratope complex which typically has a half-life 0.5 – 2 hours. The pH-dependent affinity is used routinely in the elution of antibodies at pH 2.5-3.0 during affinity chromatography^32^ suggesting the binding complex can be titrated as shown in Figure S3 for the proteins N, RBD, S(Wuhan), and S(Omicron) although the variation is complex and highly protein dependent. A linear correlation has been shown with the complex half-life and Affinity constant, K_D_ (pM) shown in Figures S4 and S5. Further, the physiological importance of lower pH is also relevant to the nasal mucosa^33^, which at pH 5.5 – 6.5, is significantly lower than the pH of the serum and the buffers used for conventional affinity measurements. A nasal mucosal protection mechanism, pH 5, would require antibody complexes to be sufficiently numerous and stable to prevent attachment to the angiotensin-converting-enzyme-2 receptor, a parameter described here as the high-affinity concentration.

## Conclusions

The immunity profile for staff in the constant-exposure cohort has been characterised in 30 dimensions: 10 variant spike proteins, total antibody concentration, AC, the high-quality antibody concertation, HQ, and the percentage of high-quality antibodies, HQ%, which are now all measures of a sterilising serum for the clearance of different variants of the SARS –CoV-2 virus. Technique independent thresholds and references ranges have been set of these serum characteristics. The patient antibody spectra show significant antibody maturation to all SARS-CoV-2 variant spike proteins, notably to the prevalent Omicron variants. Approximately, 25% of the cohort have full, pan-variant protection whereas the remainder have higher-quality antibodies focused on the preceding wave variants. A smart-boosting programme focused on high-transmission groups, such as teachers and parents/guardians of school-age children, could significantly reduce the risk of high infection levels within the population during each wave, as the pandemic transitions to endemic. Furthermore, extensive and readily measured immunity profiles may highlight specific endotype vulnerability to infection, and provide insight into long COVID complications.

## Data Availability

All data produced in the present study are available upon reasonable request to the authors

## Acknowledgements

Professor Shaw’s Research Group at Exeter University and Attomarker would like to thank the schoolteachers and staff who participated in this research project. The authors would like to thank Dr Jonathan Snicker for their guidance on the potential strategic and policy implications of the scientific findings.

## Supplementary Data

### Figures

**Figure S1.**
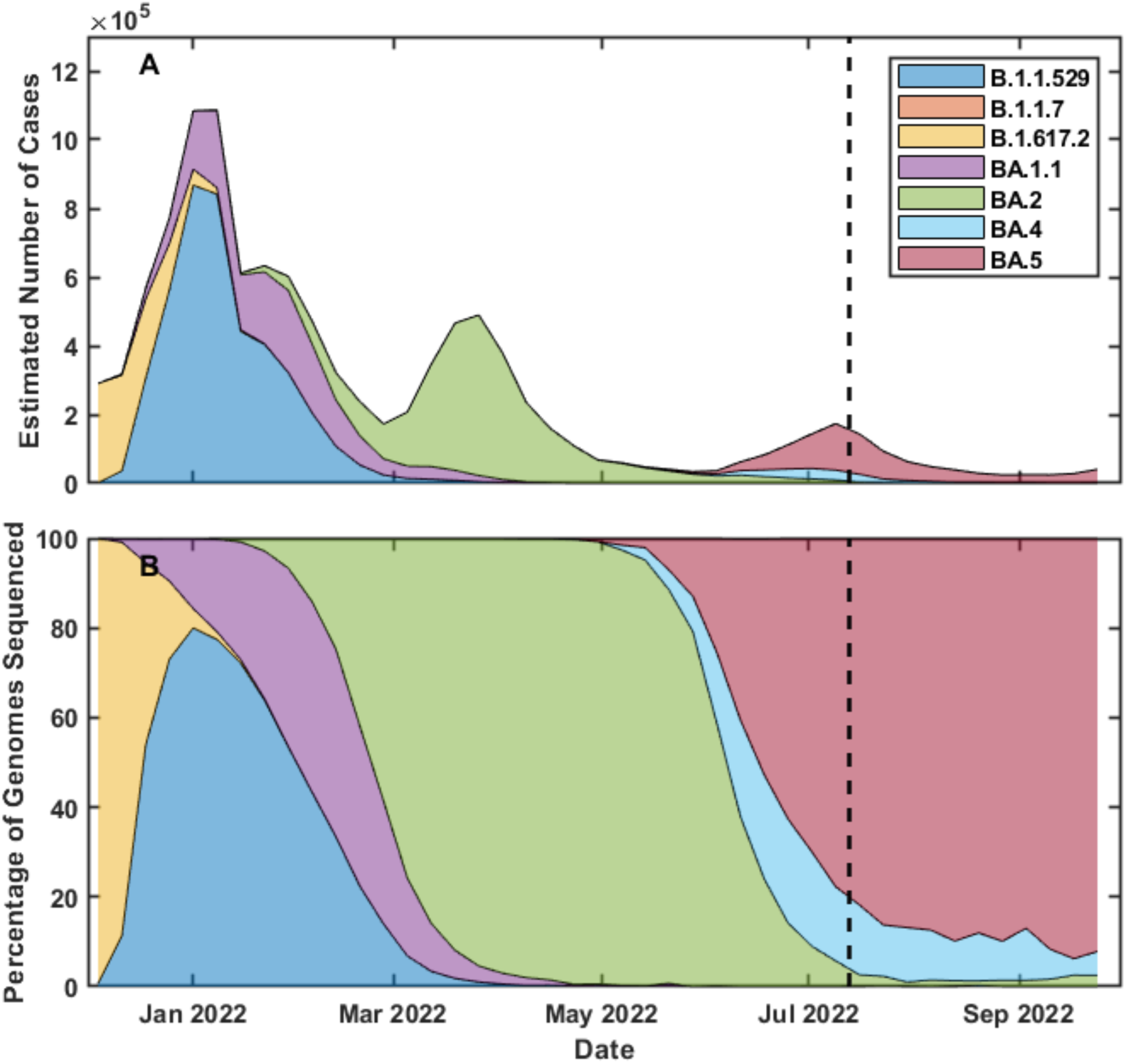
Prevalent SARS-CoV-2 variants preceding the cohort screen and the date of the cohort testing – shown using the black-dash line. Data is taken from the Wellcome Sanger Institute ^34^. – this may need to be updated.

**Figure S2.**
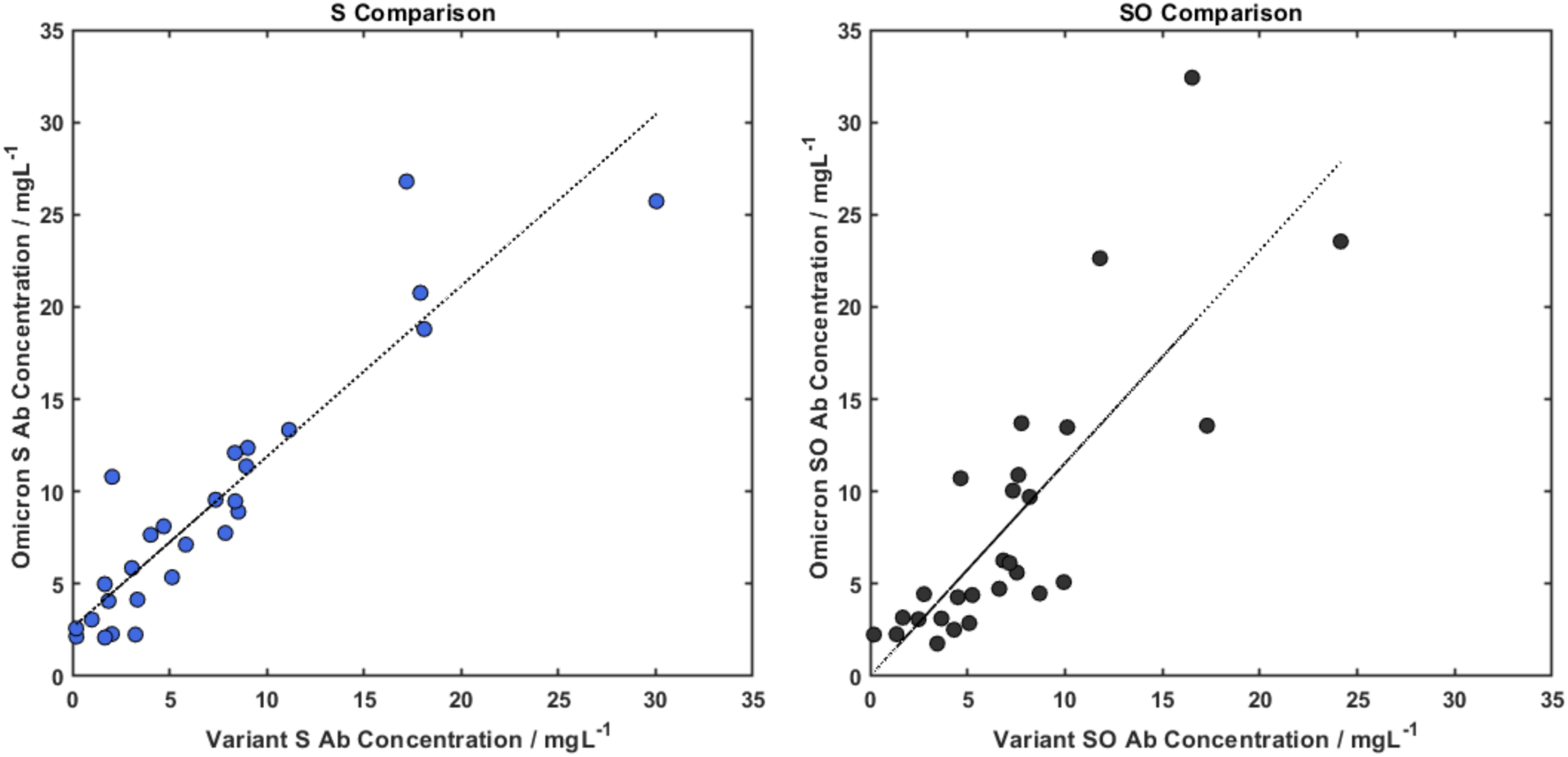
Correlation of Spike and Omicron Spike channels on the Variant and Omicron Sub-variant Arrays. – R^2^ values in the caption or the slopes and intercept.

**Figure S3.**
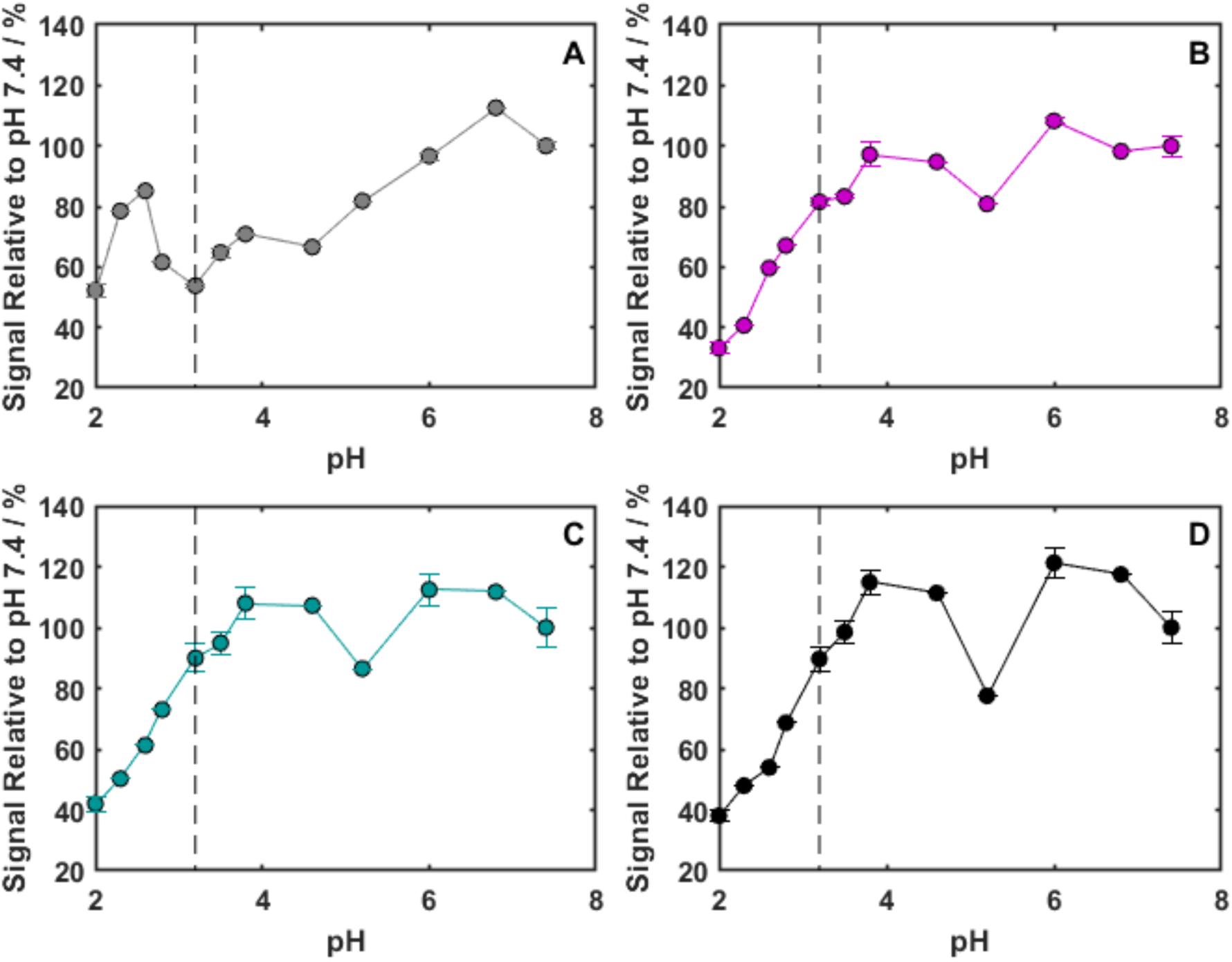
The change in concentration of the antibody assay with pH of an intermediate wash step. Protein Channels shown are the SARS-CoV-2 viral proteins: A) Nucleocapsid, B) RBD, C) S (Wuhan) and D) S (Omicron BA.1). Error bars show 1 SD. The vertical dotted line is drawn at pH 3.2.

**Figure S4.**
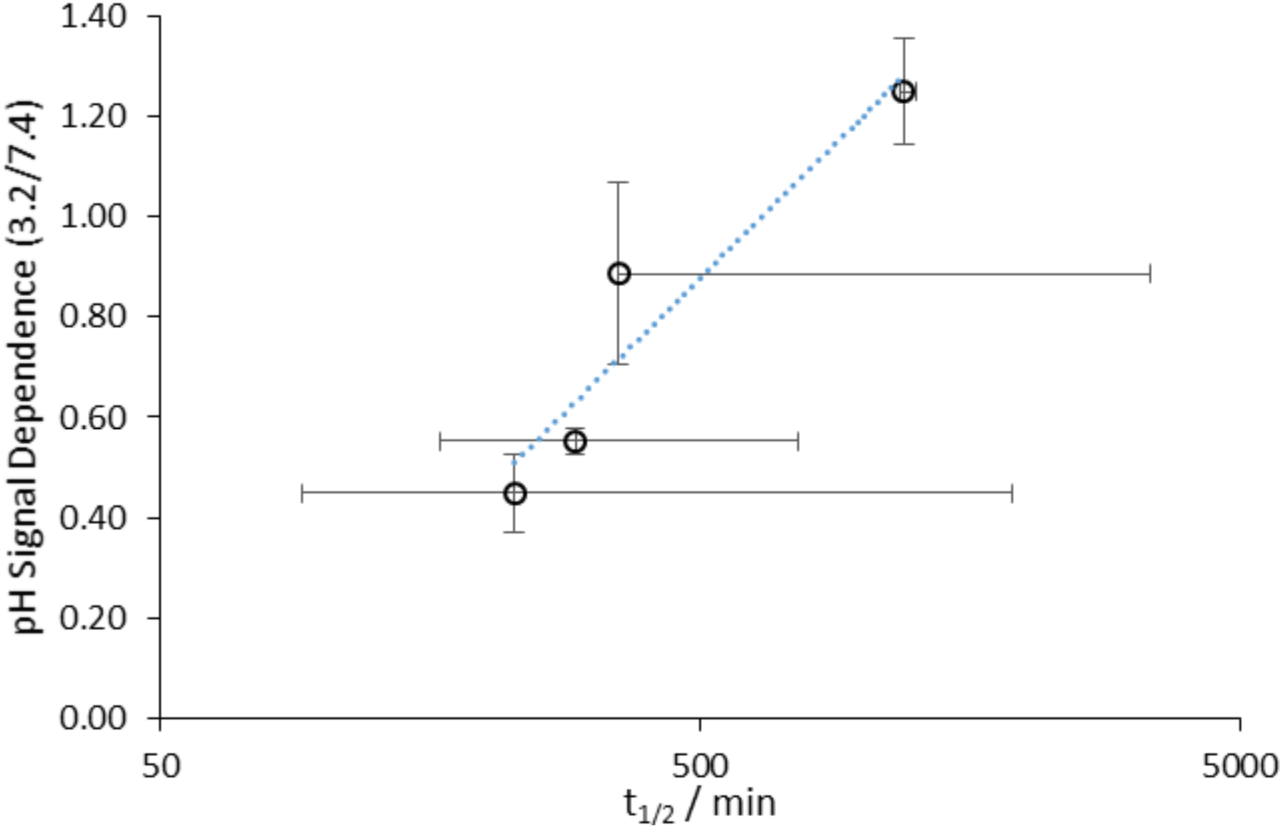
Plot of antibody-antigen half-life at pH 7.4 vs the pH dependence of the complex stability. Error bars show 1SD. The half-life was measured for four commercially sourced antibodies by recording the antibody dissociation over 20-30 minutes and fitting the data produced to a Langmuirian model. The pH dependence was determined by a sandwich assay with an intermediate wash step of 30s (between capture and detection) of varying pH.[ZHU1076 (Sigma-Aldrich), 40590-D001, 40590-T62 (Sinobiological), antiS2 1034617 (Biotechne)]. – this is going to hurt us – we need to think about it.

**Figure S5.**
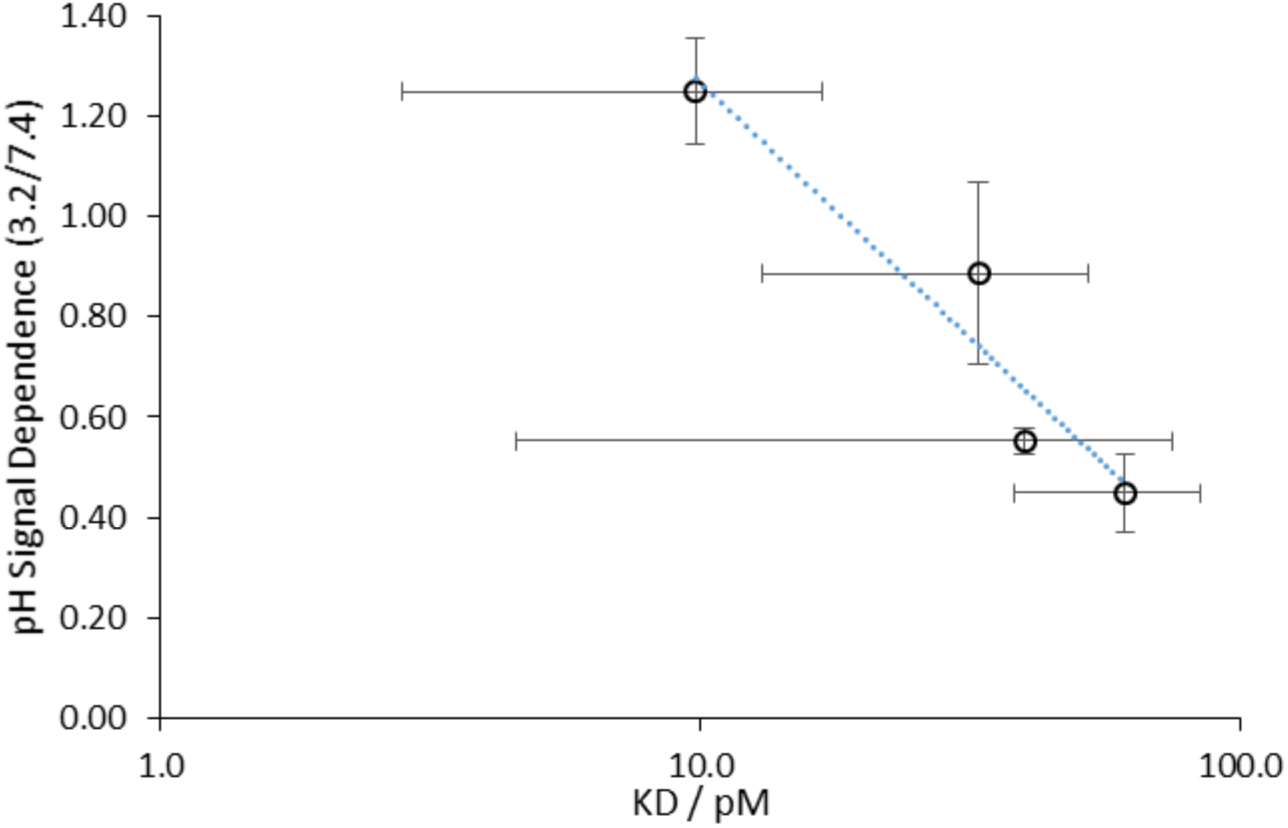
Plot of antibody K_D_ at pH 7.4 against the pH dependence of the complex stability. Error bars show 1 SD.

### Tables

**Table S1.**
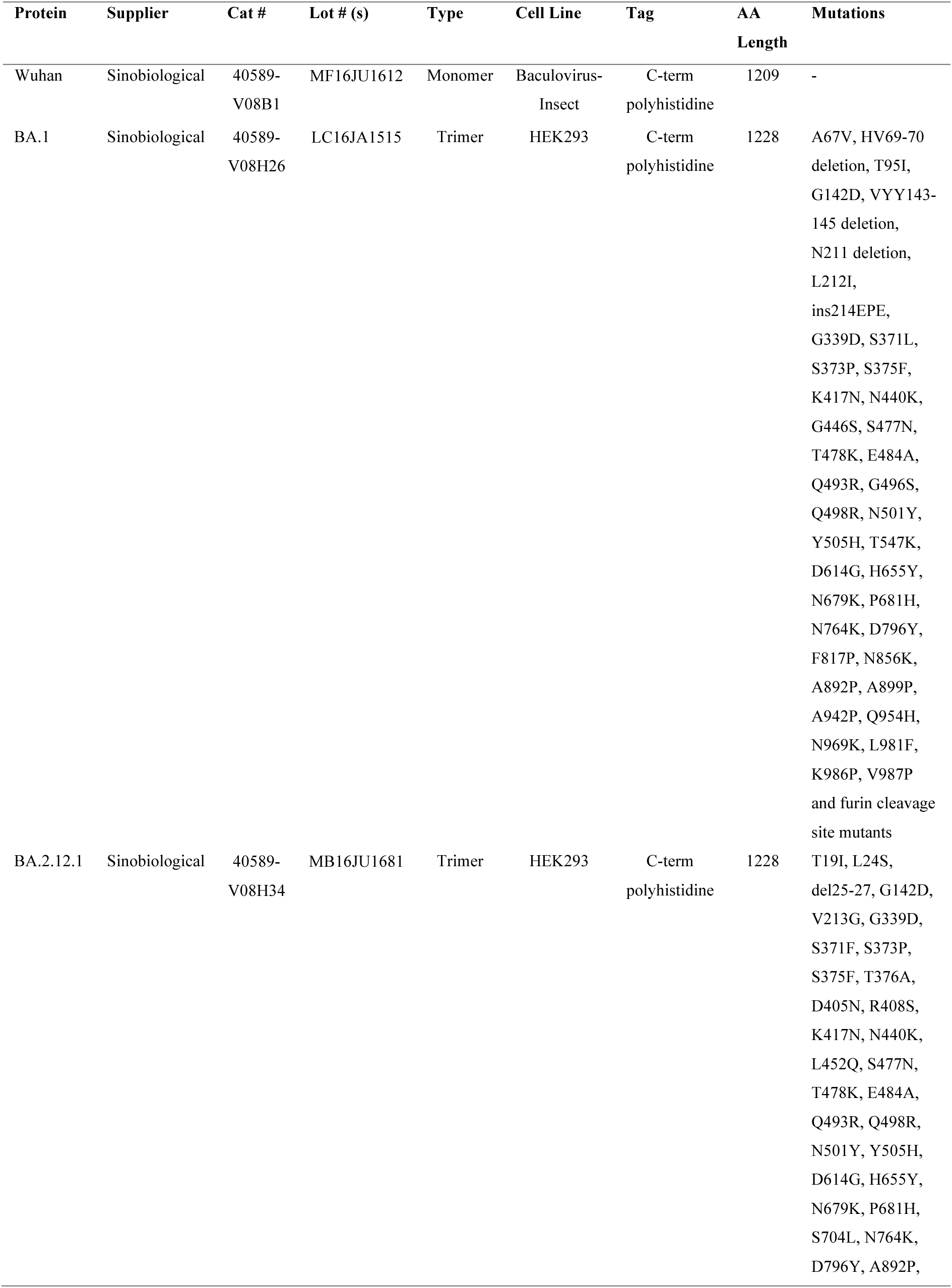
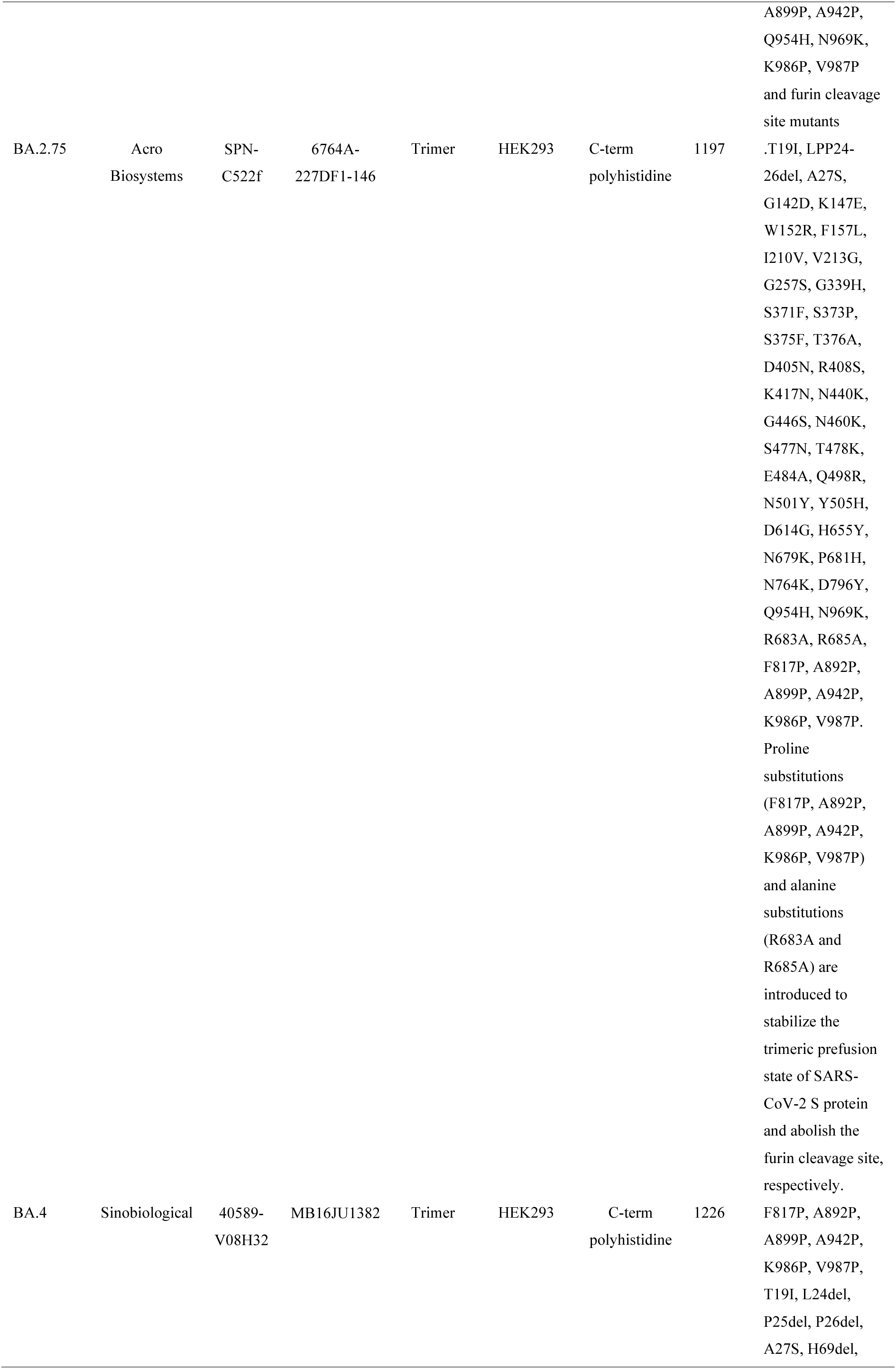
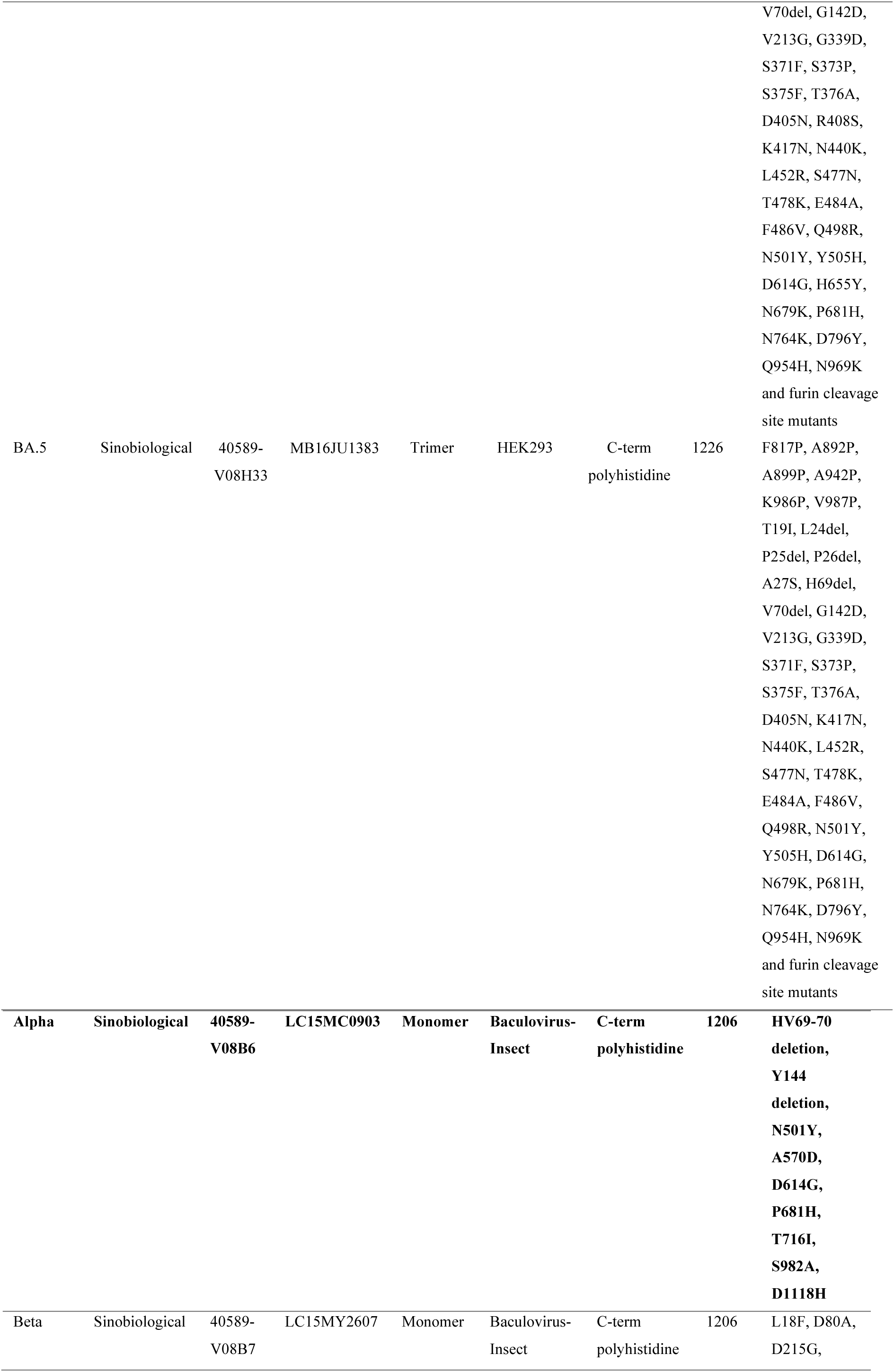
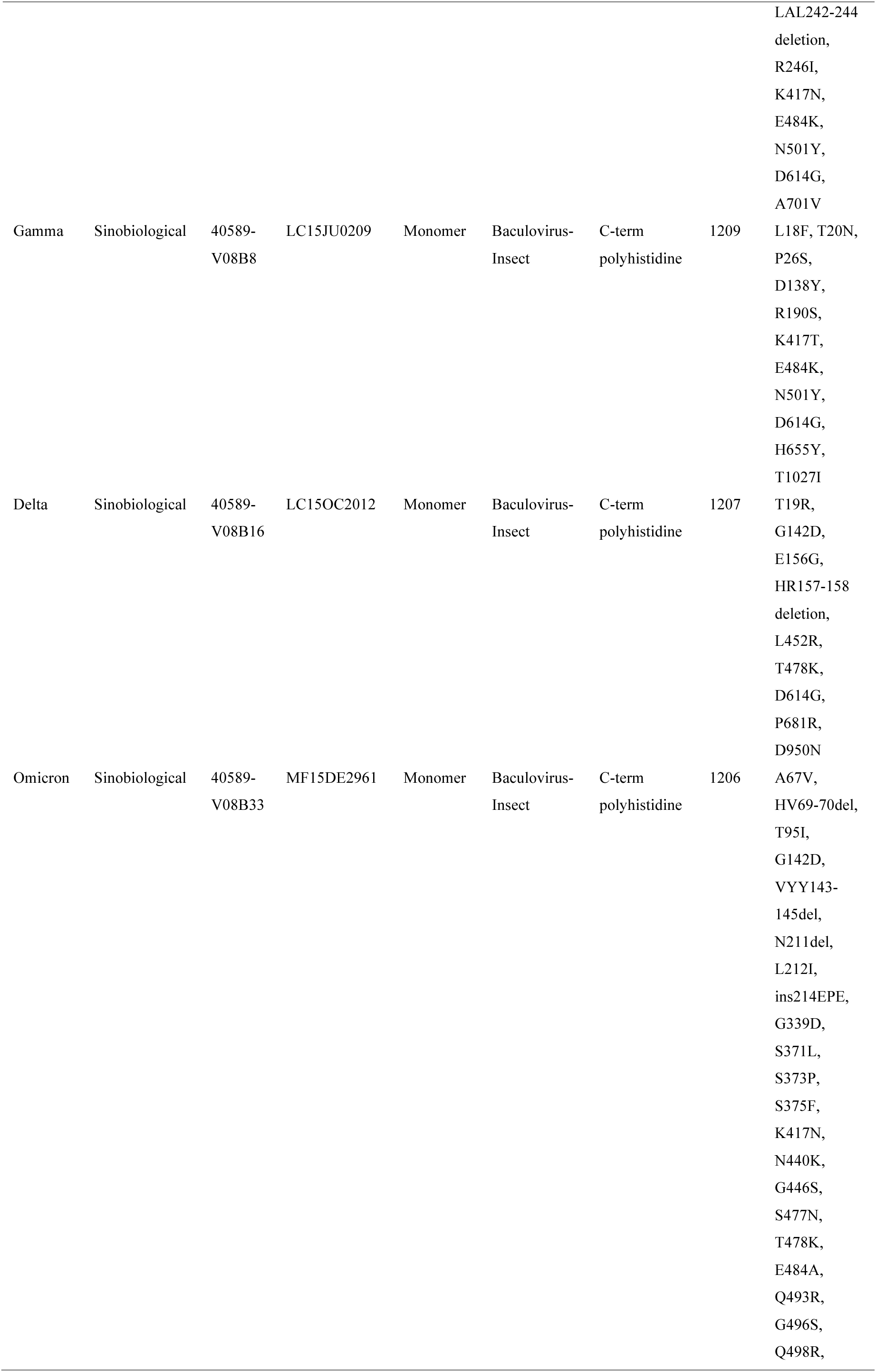
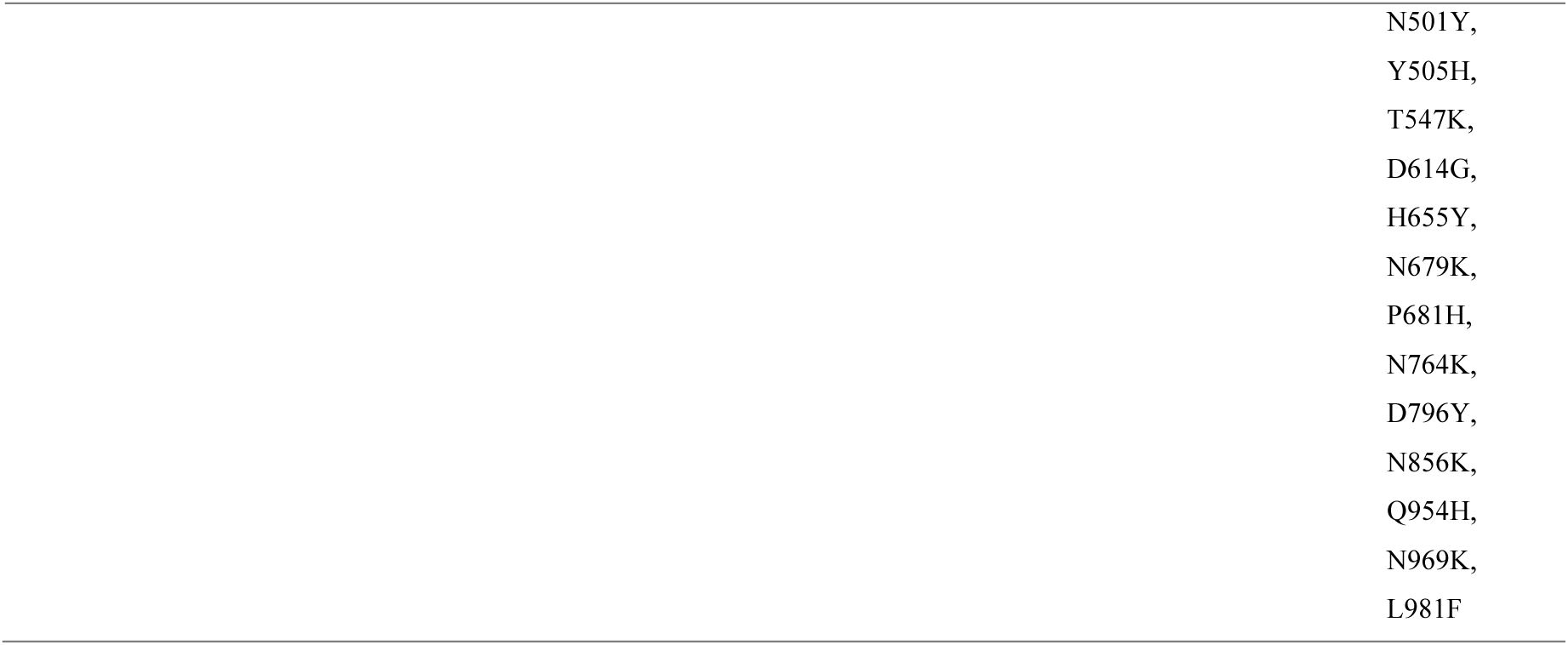
The Spike proteins used to fabricate the Omicron Sub-variant test arrays.

| Protein | Supplier | Cat # | Lot # (s) | Type | Cell Line | Tag | AA Length | Mutations |
| --- | --- | --- | --- | --- | --- | --- | --- | --- |
| Wuhan | Sinobiological | 40589-V08B1 | MF16JU1612 | Monomer | Baculovirus-Insect | C-term polyhistidine | 1209 | - |
| BA.1 | Sinobiological | 40589-V08H26 | LC16JA1515 | Trimer | HEK293 | C-term polyhistidine | 1228 | A67V, HV69-70 deletion, T95I, G142D, VYY143-145 deletion, N211 deletion, L212I, ins214EPE, G339D, S371L, S373P, S375F, K417N, N440K, G446S, S477N, T478K, E484A, Q493R, G496S, Q498R, N501Y, Y505H, T547K, D614G, H655Y, N679K, P681H, N764K, D796Y, F817P, N856K, A892P, A899P, A942P, Q954H, N969K, L981F, K986P, V987P and furin cleavage site mutants |
| BA.2.12.1 | Sinobiological | 40589-V08H34 | MB16JU1681 | Trimer | HEK293 | C-term polyhistidine | 1228 | T19I, L24S, del25-27, G142D, V213G, G339D, S371F, S373P, S375F, T376A, D405N, R408S, K417N, N440K, L452Q, S477N, T478K, E484A, Q493R, Q498R, N501Y, Y505H, D614G, H655Y, N679K, P681H, S704L, N764K, D796Y, A892P, |
|  |  |  |  |  |  |  |  | A899P, A942P, Q954H, N969K, K986P, V987P and furin cleavage site mutants |
| BA.2.75 | Acro Biosystems | SPN-C522f | 6764A-227DF1-146 | Trimer | HEK293 | C-term polyhistidine | 1197 | .T19I, LPP24-26del, A27S, G142D, K147E, W152R, F157L, I210V, V213G, G257S, G339H, S371F, S373P, S375F, T376A, D405N, R408S, K417N, N440K, G446S, N460K, S477N, T478K, E484A, Q498R, N501Y, Y505H, D614G, H655Y, N679K, P681H, N764K, D796Y, Q954H, N969K, R683A, R685A, F817P, A892P, A899P, A942P, K986P, V987P. Proline substitutions (F817P, A892P, A899P, A942P, K986P, V987P) and alanine substitutions (R683A and R685A) are introduced to stabilize the trimeric prefusion state of SARS-CoV-2 S protein and abolish the furin cleavage site, respectively. |
| BA.4 | Sinobiological | 40589-V08H32 | MB16JU1382 | Trimer | HEK293 | C-term polyhistidine | 1226 | F817P, A892P, A899P, A942P, K986P, V987P, T19I, L24del, P25del, P26del, A27S, H69del, |
|  |  |  |  |  |  |  |  | V70del, G142D,<br>V213G, G339D,<br>S371F, S373P,<br>S375F, T376A,<br>D405N, R408S,<br>K417N, N440K,<br>L452R, S477N,<br>T478K, E484A,<br>F486V, Q498R,<br>N501Y, Y505H,<br>D614G, H655Y,<br>N679K, P681H,<br>N764K, D796Y,<br>Q954H, N969K<br>and furin cleavage<br>site mutants |
| BA.5 | Sinobiological | 40589-V08H33 | MB16JU1383 | Trimer | HEK293 | C-term<br>polyhistidine | 1226 | F817P, A892P,<br>A899P, A942P,<br>K986P, V987P,<br>T19I, L24del,<br>P25del, P26del,<br>A27S, H69del,<br>V70del, G142D,<br>V213G, G339D,<br>S371F, S373P,<br>S375F, T376A,<br>D405N, K417N,<br>N440K, L452R,<br>S477N, T478K,<br>E484A, F486V,<br>Q498R, N501Y,<br>Y505H, D614G,<br>N679K, P681H,<br>N764K, D796Y,<br>Q954H, N969K<br>and furin cleavage<br>site mutants |
| **Alpha** | **Sinobiological** | **40589-**<br>**V08B6** | **LC15MC0903** | **Monomer** | **Baculovirus-**<br>**Insect** | **C-term**<br>**polyhistidine** | **1206** | **HV69-70**<br>**deletion,**<br>**Y144**<br>**deletion,**<br>**N501Y,**<br>**A570D,**<br>**D614G,**<br>**P681H,**<br>**T716I,**<br>**S982A,**<br>**D1118H** |
| Beta | Sinobiological | 40589-V08B7 | LC15MY2607 | Monomer | Baculovirus-Insect | C-term<br>polyhistidine | 1206 | L18F, D80A,<br>D215G, |
|  |  |  |  |  |  |  |  | LAL242-244<br>deletion,<br>R246I,<br>K417N,<br>E484K,<br>N501Y,<br>D614G,<br>A701V |
| Gamma | Sinobiological | 40589-V08B8 | LC15JU0209 | Monomer | Baculovirus-Insect | C-term<br>polyhistidine | 1209 | L18F, T20N,<br>P26S,<br>D138Y,<br>R190S,<br>K417T,<br>E484K,<br>N501Y,<br>D614G,<br>H655Y,<br>T1027I |
| Delta | Sinobiological | 40589-V08B16 | LC15OC2012 | Monomer | Baculovirus-Insect | C-term<br>polyhistidine | 1207 | T19R,<br>G142D,<br>E156G,<br>HR157-158<br>deletion,<br>L452R,<br>T478K,<br>D614G,<br>P681R,<br>D950N |
| Omicron | Sinobiological | 40589-V08B33 | MF15DE2961 | Monomer | Baculovirus-Insect | C-term<br>polyhistidine | 1206 | A67V,<br>HV69-70del,<br>T95I,<br>G142D,<br>VYY143-145del,<br>N211del,<br>L212I,<br>ins214EPE,<br>G339D,<br>S371L,<br>S373P,<br>S375F,<br>K417N,<br>N440K,<br>G446S,<br>S477N,<br>T478K,<br>E484A,<br>Q493R,<br>G496S,<br>Q498R, |

|  |
| --- |
| N501Y,<br>Y505H,<br>T547K,<br>D614G,<br>H655Y,<br>N679K,<br>P681H,<br>N764K,<br>D796Y,<br>N856K,<br>Q954H,<br>N969K,<br>L981F |

**Table S2.** Gradient (m), intercept (c) and R^2^ values for the correlation of similar channels on the Spike Variant Array and the Spike Omicron Array.

| Protein Pair x/y | Slope | Intercept | $R^2$ |
| --- | --- | --- | --- |
| S (monomer)/S (monomer) | 0.93 | 2.6 | 0.92 |
| SO (monomer) /SO (trimer) | 1.15 | 0.0 | 0.81 |

**Table S4.**
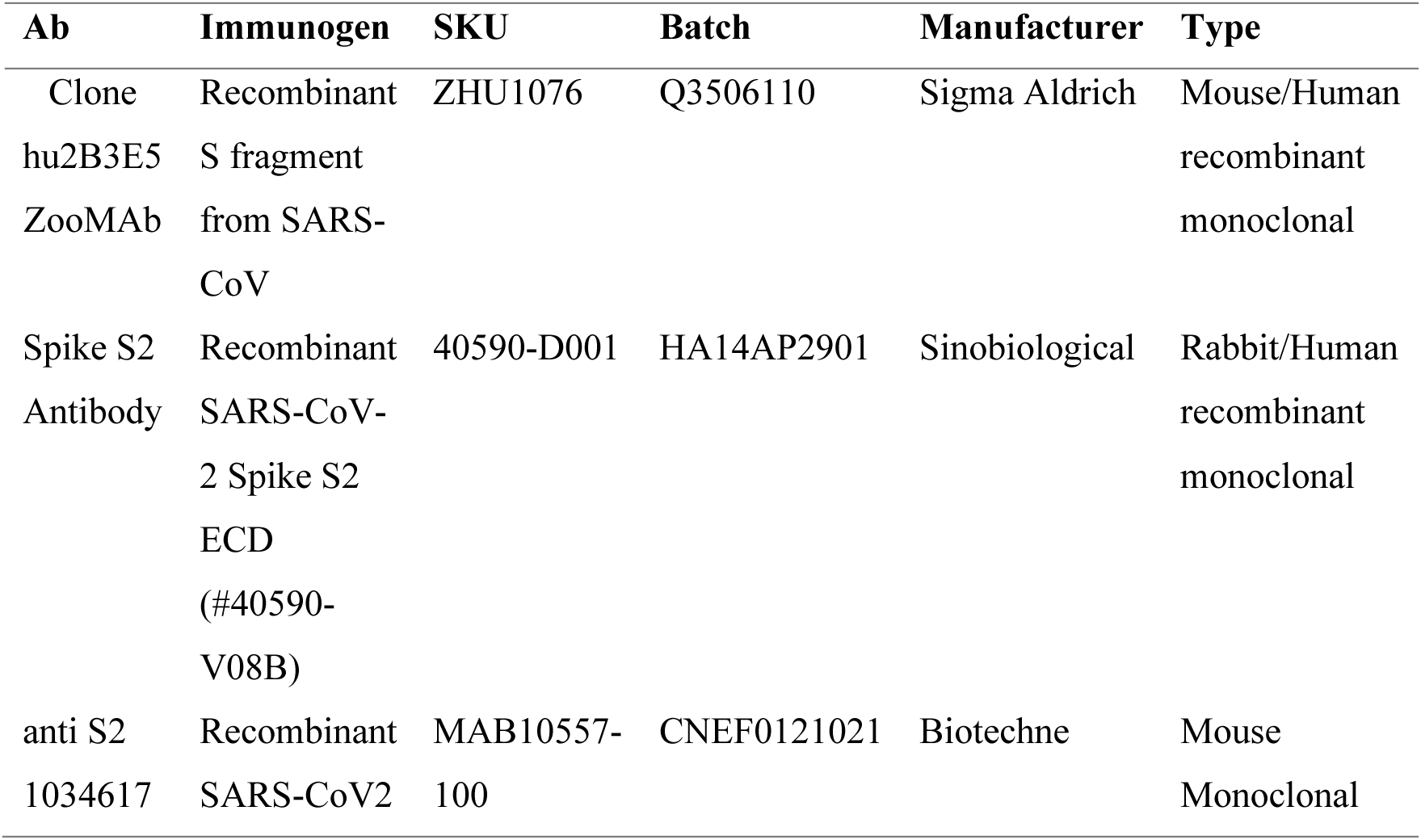
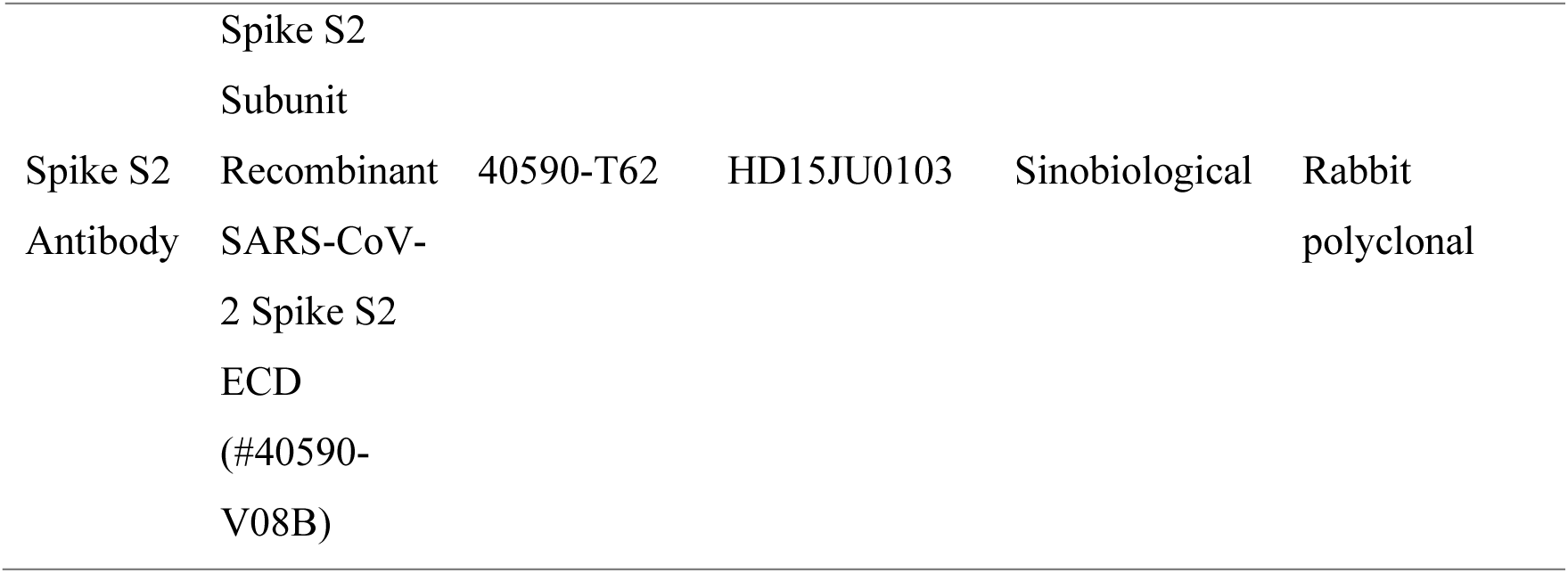
Antibodies used for production of t_1/2_ to k_d_ plot.

**Table S5.** Ranges for the AC, HQ and HQ% derived from the cohort showing median, d Lower Quartile (LQ) and Upper Quartile (UQ: HQ% W, BA.1, BA2.12.1 BA.2.75. BA.4 and BA.5.

|  | Wuhan | Alpha | Beta | Gamma | Delta | BA.1 | BA.2.12.1 | BA.2.75 | BA.4 | BA.5 |
| --- | --- | --- | --- | --- | --- | --- | --- | --- | --- | --- |
| **AC / mg L<sup>-1</sup>** |  |  |  |  |  |  |  |  |  |  |
| **LQ** | 3.2 | 5.3 | 3.7 | 5.7 | 2.5 | 3.5 | 6.9 | 5.1 | 6.2 | 5.4 |
| **Median** | 6.4 | 8.6 | 8.8 | 10.2 | 6.5 | 6.6 | 8.8 | 9.9 | 9.9 | 8.2 |
| **UQ** | 10.2 | 11.3 | 13.1 | 12.8 | 11.0 | 9.1 | 14.4 | 14.5 | 14.0 | 13.0 |
| **HQ / mg L<sup>-1</sup>** |  |  |  |  |  |  |  |  |  |  |
| **LQ** | 1.0 | 0.6 | 0.3 | 0.4 | 0.9 | 1.5 | 3.2 | 3.7 | 2.8 | 2.7 |
| **Median** | 2.6 | 2.3 | 0.6 | 1.2 | 1.7 | 2.2 | 4.8 | 5.7 | 3.6 | 3.4 |
| **LQ** | 1.0 | 0.6 | 0.3 | 0.4 | 0.9 | 1.5 | 3.2 | 3.7 | 2.8 | 2.7 |
| **HQ / %** |  |  |  |  |  |  |  |  |  |  |
| **LQ** | 32 | 10 | 5 | 5 | 22 | 34 | 49 | 47 | 37 | 37 |
| **Median** | 44 | 27 | 15 | 11 | 32 | 43 | 53 | 58 | 39 | 42 |
| **UQ** | 51 | 43 | 20 | 32 | 48 | 54 | 63 | 68 | 47 | 54 |
| **Reference Range Mean and IQR** |  |  |  |  |  |  |  |  |  |  |
| **49% (32% – 68%)** |  |  |  |  |  |  |  |  |  |  |

**Table S2.**
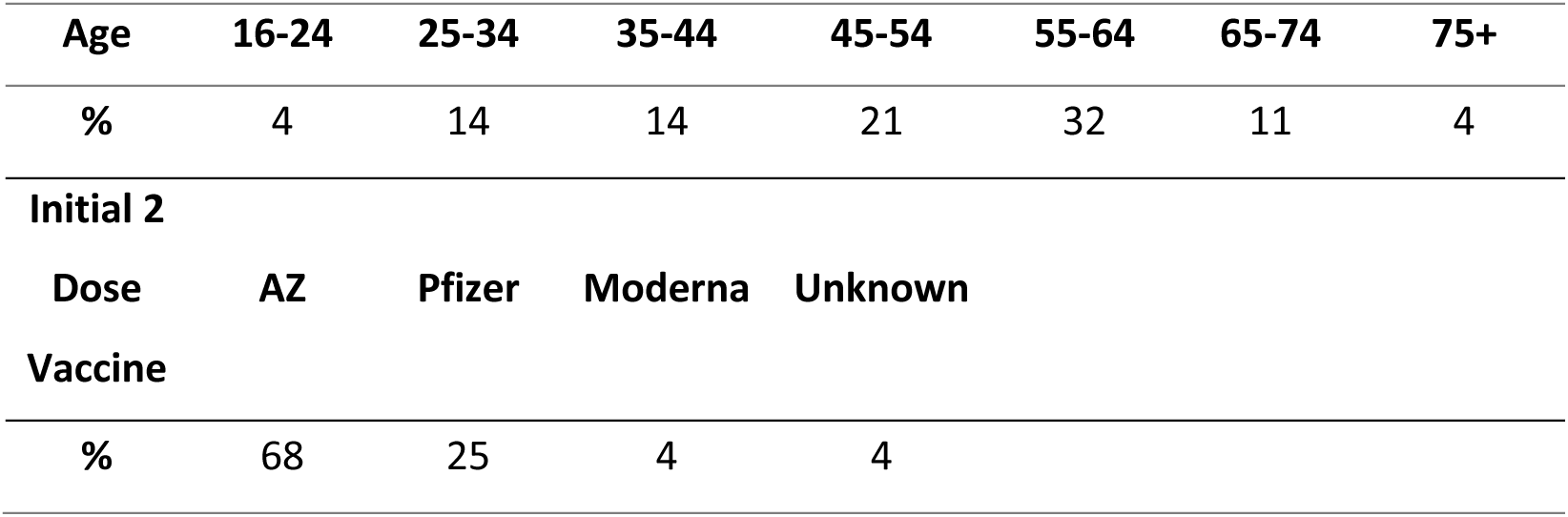
Additional cohort demographics.

| Age | 16-24 | 25-34 | 35-44 | 45-54 | 55-64 | 65-74 | 75+ |
| --- | --- | --- | --- | --- | --- | --- | --- |
| % | 4 | 14 | 14 | 21 | 32 | 11 | 4 |
| Initial 2 |  |  |  |  |  |  |  |
| Dose | AZ | Pfizer | Moderna | Unknown |  |  |  |
| Vaccine |  |  |  |  |  |  |  |
| % | 68 | 25 | 4 | 4 |  |  |  |

**Table S3.** Channels grouped according to time. Early = all Pre-Omicron.

|  | Early Variants | Omicron Variants |
| --- | --- | --- |
| **AC / mg L<sup>-1</sup>** |  |  |
| LQ | 4.4 | 5.3 |
| Median | 7.8 | 8.2 |
| UQ | 12.2 | 13.7 |
| **HQ / mg L<sup>-1</sup>** |  |  |
| LQ | 0.5 | 2.6 |
| Median | 1.7 | 4.0 |
| LQ | 4.5 | 7.5 |
| **HQ / %** |  |  |
| LQ | 11 | 38 |
| Median | 27 | 47 |
| UQ | 45 | 58 |

## Related Publications

Paper 1 – Mass-Standardised Quantitative Measurements of the Antibody Levels for SARS-CoV-2 beyond Correlates of Protection and Clearance

Paper 2 – Mass-Standardised Differential Antibody Binding to a Spectrum of SARS-CoV-2 Variant Spike Proteins: Wuhan, Alpha, Beta, Gamma, Delta, Omicron BA.1, BA.4/5, BA.2.75 and BA.2.12.1 variants – Antibody Immunity Endotypes

Paper 3 – this paper – Mass-Standardised Antibody Affinity Maturation to the Spike Protein of SARS-CoV-2 Omicron Variants in a Teachers Cohort: forgiving Original Antigenic Sin

Paper 4 – Diagnostic Classification for Long Covid Patients identifying Persistent Virus and Hyperimmune Pathophysiologies leading to a Test-and-Treat Protocol

